# Temporal course of SARS-CoV-2 antibody positivity in patients with COVID-19 following the first clinical presentation

**DOI:** 10.1101/2020.10.17.20214445

**Authors:** Martin Risch, Myriam Weber, Sarah Thiel, Kirsten Grossmann, Nadia Wohlwend, Thomas Lung, Dorothea Hillmann, Michael Ritzler, Francesca Ferrara, Susanna Bigler, Konrad Egli, Thomas Bodmer, Mauro Imperiali, Yacir Salimi, Felix Fleisch, Alexia Cusini, Harald Renz, Philipp Kohler, Pietro Vernazza, Christian Kahlert, Matthias Paprotny, Lorenz Risch

**Affiliations:** Zentrallabor, Kantonsspital Graubünden, Loësstrasse 170, 7000 Chur, Switzerland; Liechtensteinisches Landesspital, Heiligkreuz, 9490 Vaduz, Liechtenstein; Labormedizinisches zentrum Dr. Risch, Wuhrstrasse 14, 9490 Vaduz, Liechtenstein; Private Universität im Fürstentum Liechtenstein, Dorfstrasse, 9495 Triesen Liechtenstein; Labormedizinisches zentrum Dr. Risch, Waldeggstrasse 37, 3097 Liebefeld, Switzerland; Centro medicina di laboratorio Dr. Risch, Via Arbostra 2, 6963 Pregassona, Switzerland; Clm Dr Risch arc lémanique SA, Chemin de l’Esparcette 10, 1023 Crissier, Switzerland; Division of Infectious Diseases, Cantonal Hospital Chur, Loësstrasse 170, 7000 Chur, Switzerland; Institute of Laboratory Medicine and Pathobiochemistry, Molecular Diagnostics, Philipps University Marburg, University Hospital Giessen and Marburg, Baldingerstraße, 35043 Marburg, Germany; Cantonal Hospital St Gallen, Department of Infectious Diseases and Hospital Epidemiology, Rohrschacherstrasse 95 9007 St Gallen, Switzerland; Children’s Hospital of Eastern Switzerland, Department of Infectious Diseases and Hospital Epidemiology, Claudiusstrasse 6, 9006 St. Gallen, Switzerland; Center of Laboratory Medicine, University Institute of Clinical Chemistry, University of Bern, Inselspital, 3010 Bern, Switzerland

**Author notes:** Correspondence to: Prof. Dr. med. Lorenz Risch, MPH, MHA, Labormedizinisches zentrum Dr. Risch, Wuhrstrasse 14, 9490 Vaduz.

**Keywords:** Antibody, Coronavirus, COVID-19, Kinetics, Predictive Values, SARS-CoV-2, Sensitivity, Serology

## Abstract

Knowledge of the sensitivities of severe acute respiratory syndrome coronavirus-2 (SARS-CoV-2) antibody tests beyond 35 days after the clinical onset of COVID-19 is insufficient. We aimed to describe positivity rate of SARS-CoV-2 assays employing three different measurement principles over a prolonged period. Two hundred sixty-eight samples from 180 symptomatic patients with COVID-19 and a reverse transcription polymerase chain reaction (RT-PCR) test followed by serological investigation of SARS-CoV-2 antibodies were included.. We conducted three chemiluminescence (including electrochemiluminscence, ECLIA), four enzyme linked immunosorbent assay (ELISA), and one lateral flow immunoassay (LFIA) test formats. Positivity rates, as well as positive (PPV) and negative predictive values (NPV) were calculated for each week after the first clinical presentation for COVID-19. Furthermore, combinations of tests were assessed within an orthogonal testing approach employing two independent assays and predictive values were calculated. Heat maps were constructed to graphically illustrate operational test characteristics. During a follow-up period of more than 9 weeks, chemiluminescence assays and one ELISA IgG test showed stable positivity rates after the third week. With the exception of ECLIA, the PPVs of the other chemiluminescence assays were ≥95% for COVID-19 only after the second week. ELISA and LFIA had somewhat lower PPVs. IgM exhibited insufficient predictive characteristics. An orthogonal testing approach provided PPVs ≥95% for patients with a moderate pretest probability (e.g., symptomatic patients), even for tests with a low single test performance. After the second week, NPVs of all but IgM assays were ≥95% for patients with low to moderate pretest probability. The confirmation of negative results using an orthogonal algorithm with another assay provided lower NPVs than the single assays. When interpreting results from SARS-CoV-2 tests, the pretest probability, time of blood draw and assay characteristics must be carefully considered. An orthogonal testing approach increases the accuracy of positive, but not negative, predictions.

## Introduction

Coronavirus Disease 2019 (COVID-19) is a pandemic that has challenged healthcare systems worldwide ^1^. The disease is caused by the severe acute respiratory syndrome coronavirus 2 (SARS-CoV-2) ^2^. Its diagnosis is based on clinical symptoms and signs, radiological imaging, and the detection of the virus with reverse transcription polymerase chain reaction (RT-PCR) or antigen testing mainly in respiratory specimens, and serological confirmation of SARS-CoV-2-specific antibodies in serum ^3,4^. Although RT-PCR has been reported to possess 100% specificity, it does not provide a 100% sensitivity in diagnosing COVID-19 ^5^, due to issues related to the preanalytical sample quality as well as local and temporal changes in viral shedding ^5-7^. In the clinic, serological testing has increasingly become important to clarify RT-PCR-negative patients with a high clinical suspicion of COVID-19 (“false negatives”) ^8^. Furthermore, serological testing has also become important for surveillance and tracing purposes to identify the disease prevalence in a distinct population or identify close contacts of patients with asymptomatic COVID-19 ^9^.

Recently, a Cochrane review systematically analyzed the sensitivity of different assays in diagnosing COVID-19 and concluded that the time since the onset of symptoms is a critical determinant of test sensitivity ^3^. Antibody tests can play a useful role after the first week of symptom onset ^3^. However, the authors obtained very little data to describe the sensitivity of the different tests after 35 days. The present study aimed to describe the positivity rates of different serological tests after a diagnosis of COVID-19 for a prolonged period of up to 9 weeks.

## Methods

### Study setting and study population

This retrospective study was conducted using anonymized samples. These samples were obtained from patients in Switzerland and the principality of Liechtenstein and sent to a binational group of medical laboratories (labormedizinische zentren Dr. Risch, Switzerland and Liechtenstein). Samples were included in the analysis when patients had a positive RT-PCR for COVID-19 prior to serum sampling. The vast majority of the RT-PCR-positive results was obtained during March and April 2020, and three further RT-PCR-positive results were obtained until June 12, 2020. The serum samples mainly originated from outpatient settings. During March and April, RT-PCR testing in Switzerland and Liechtenstein was offered to individuals presenting with a fever of 38°C and respiratory symptoms, whereas after April 2020, testing was offered to individuals displaying any possible COVID-19 symptom ^10^. The study protocol was verified by the cantonal ethics boards of Zurich (BASEC Req-20-00587) and Eastern Switzerland (EKOS; BASEC Nr. Req-2020-00586). Informed consent for performing a laboratory analysis of anonymized samples was waived.

### Laboratory methods

For each serum sample, the age and sex of the individual, the type of clinical setting and the number of days from sample collection to RT-PCR were available. Serum employed for testing was either freshly collected or stored for less than 3 months at −25°C. For antibody testing using the chemiluminescence technique, antibodies were tested with three chemiluminescence assays, four enzyme linked immunosorbent assays (ELISA), and one lateral flow immunoassay, as detailed in table 1. The manufacturers’ cut-off values as well as the imprecision of the assays are also shown in table 1: COI (cut-off index) for electrochemiluminescence assay (ECLIA), and S/C (S/C=extinction of the patient sample divided by the extinction of the calibrator) for the other chemiluminescence (i.e. CMIA, chemiluminescent microparticle immunoassay, and LIA, Luminescence immunoassay) and ELISA assays. All tests were performed and analyzed according to the manufacturer’s instructions. All assays were CE-marked. Lab technicians reading the lateral flow test format were not aware of the RT-PCR result or the results of other antibody tests when manually reading the lateral flow test cassettes.

**Table 1.** Employed tests for determination of anti-SARS-CoV-2 antibodies. ELISA=enzyme linked immunosorbent assay; Ig= total immunoglobulin; N= nucleocapsid antigen; S1= spike protein S1 subunit: S1/S2= spike protein S1/S2 subunits; CV=coefficient of variation. DSX is an ELISA analyzer (Dynex Technologies, Denkendorf, Germany).

| Test Acronym | Measurement principle | Test name | Manufacturer | Analyzer | Isotype | Target antigen | Cut-off | CV (%) |
| --- | --- | --- | --- | --- | --- | --- | --- | --- |
| CMIA | Chemiluminescence microparticle immunoassay | SARS-CoV-2 IgG | Abbott Diagnostics, Baar, Switzerland | Architect i2000 | IgG | N | $\geq 1.4$ | 3.6 |
| ECLIA | Electrochemiluminescence immunoassay | Elecsys® Anti-SARS-CoV-2 | Roche Diagnostics, Rotkreuz, Switzerland | COBAS 6000 | Total Ig | N | $> 1$ | 2.7 |
| LIA | Luminescence immunoassay | LIAISON® SARS-CoV-2 S1/S2 IgG | Diasorin, Luzern, Switzerland | Liaison | IgG | S1/S2 | $> 12$ | 5.4 |
| EI IgG | ELISA | EUROIMMUN Anti-SARS-CoV-2 ELISA IgG (EI IgG) | Euroimmun, Luzern, Switzerland | DSX | IgG | S1 | $\geq 1.1$ | 3.6 |
| EI IgA | ELISA | EUROIMMUN Anti- | Euroimmun, Luzern, | DSX | IgA | S1 | $\geq 1.1$ | 4.6 |
|  |  | SARS-CoV-2 ELISA<br>IgG (EI IgA) | Switzerland |  |  |  |  |  |
| EDI IgG | ELISA | EDI™ Novel<br>Coronavirus COVID-19 IgG | Epitope<br>Diagnostics, Inc.,<br>San Diego, USA | DSX | IgG | N | <u>Manufacturer</u><br><u>specific</u> | 11.2 |
| EDI IgM | ELISA | EDI™ Novel<br>Coronavirus COVID-19 IgM | Epitope<br>Diagnostics, Inc.,<br>San Diego, USA | DSX | IgM | N | <u>Manufacturer</u><br><u>specific</u> | 9.7 |
| LFIA | Lateral flow immunoassay | SGTi-flex COVID-19<br>IgM/IgG | Sugentech Inc,<br>Daejeon, South<br>Korea | N/A | IgG/IgM | N | Qualitative<br>reading | N/A |

### Statistical methods

Continuous variables are presented as medians and interquartile ranges [IQRs], whereas proportions are presented as percentages and 95% confidence intervals (CIs). The positive rates of the different assays were calculated as fractions of positive results of all investigated samples from patients with positive RT-PCR results (i.e., sensitivity) at different weekly time periods after the first clinical presentation for COVID-19. Statistically significant differences in proportions were tested using the Chi-square test. Furthermore, operative test characteristics stratified according to the time point of blood sampling, and pretest probabilities were assessed by calculating negative (NPV) and positive (PPV) predictive values ^11^. For this analysis, the following specificity values, as evaluated in another cohort investigated by some of the authors of this report in a cohort of 1002 healthcareworkers without COVID-19 ^12^, were employed: 99.5% for the CMIA, 99.7% for the LIA, 99.9% for the ECLIA, 99.2% for the EI IgG ELISA, 99.2% for the EI IgA ELISA, 95.8% for the EDI IgG ELISA, 96.7% for the EDI IgM ELISA, 99.5% for the IgG LFIA, 95.6% for the IgM LFIA, and 95.2% for IgG and/or IgM in the LFIA. For an illustration of predictive values, heat maps were constructed to graphically display predictive values stratified according to the time point of blood sampling and pretest probability. Pretest probabilities were considered very low (1%; e.g., asymptomatic persons in very low prevalence regions), low (<10%; e.g., asymptomatic persons in low prevalence regions), moderate (10-25%; e.g., symptomatic persons with a suspicion of COVID-19) and high (≥25%; e.g., symptomatic household contacts of patients with COVID-19) 10. In addition to the predictive values for single tests, we also evaluated orthogonal testing algorithms that sequentially combined the test characteristics of two tests according to the method proposed by the Food and Drug Administration (FDA) and the Centers of Disease Control (CDC) in the United States of America ^13,14^. Predictive values were rounded to integers. P-values <0.05 were considered statistically significant. Statistical computations were performed with Medcalc version 18.11.3 (Mariakerke, Belgium). Graphs were drawn with Microsoft Excel 2016 MSO (16.0.8431.2046) (Microsoft Inc., Seattle, USA) using the linear interpolation function.

## Results

### Sensitivity

Two hundred sixty-eight samples from 180 patients were available. The median age at diagnosis was 56 years IQR [38,72]. Not all samples were tested with all tests. The number of tested samples and the number of positive samples is presented in table 2. The rates of positive samples over the different weeks are illustrated in Figure 1. Figure 1a shows the positivity rates of the chemiluminescence assays, and all three investigated formats had the highest positivity rate during the 3^rd^ week after the clinical presentation. After this period, the positivity rates for all three investigated assays decreased to approximately 92% and remain stable. For the ELISAs, a peak was also observed at week 3 for IgG and IgM, whereas the peak for IgA occurred at week 5. IgG exhibited a different behavior, according to the test manufacturer. Although the EI positivity rate remained constant after 9 weeks, the positivity rate of EDI decreased by approximately 40%. The IgM positivity rate decreased even further to approximately 15%. IgA also showed a decrease in the positivity rate to approximately 80%. In the LFIA, IgM showed an earlier increase in positivity rates than IgG, with IgG and IgM positivity rates peaking at a value of approximately 80% at week 3. Notably, the LFIA displayed a significantly higher sensitivity than the LIA, the EI IgG ELISA, and the EDI IgM ELISA (78 vs. 50%, p=0.02; vs. 36%, p=0.004; vs. 53%; p=0.04) at week 2. The positivity rate of IgM decreased to 30% after 9 weeks, whereas the IgG positivity rate decreased to 70%. The LFIA test format reporting combined reaction of both IgG and IgM showed a positivity rate of approximately 90% at week 3, which decreased to somewhat lower values at week 9.

**Table 2.**
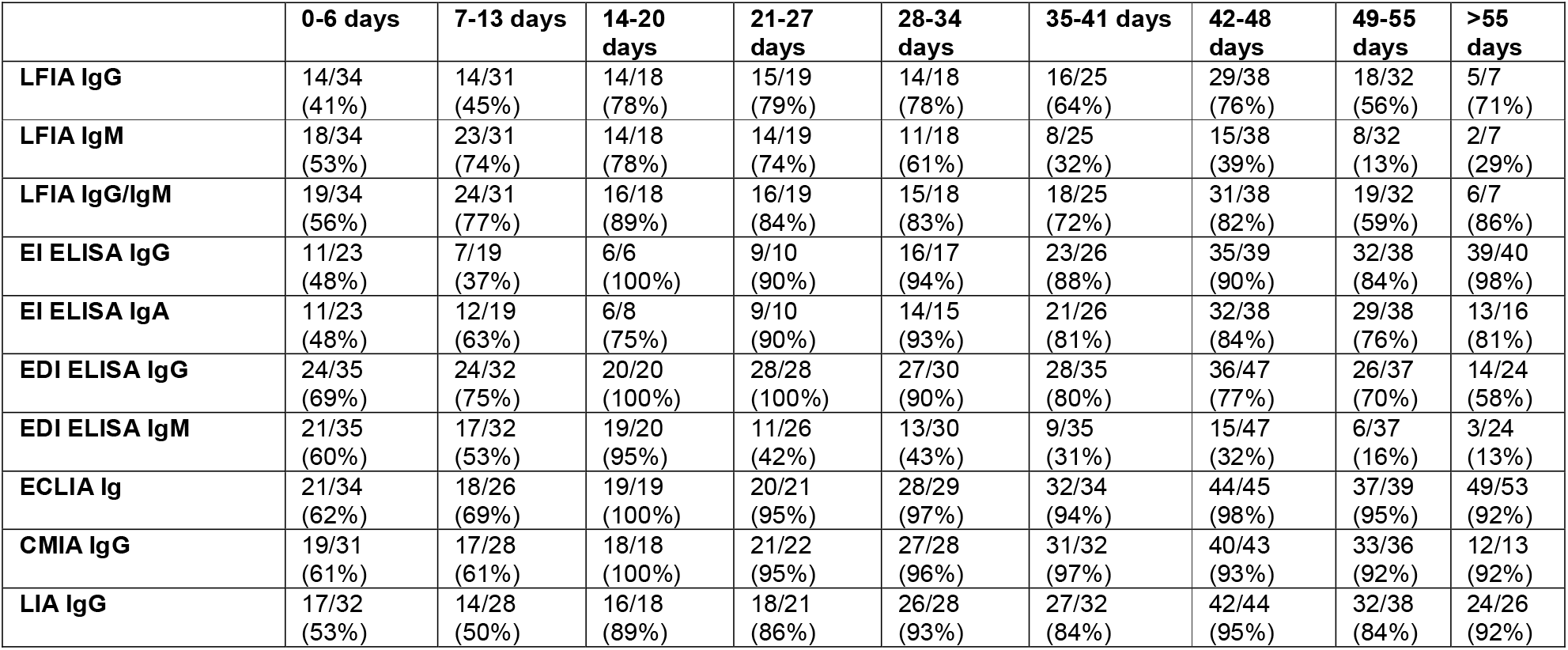
Positive samples among all investigated samples from patients with COVID-19 stratified according to time of serum sampling since the first clinical presentation. Positivity rates are given in brackets.

|  | 0-6 days | 7-13 days | 14-20 days | 21-27 days | 28-34 days | 35-41 days | 42-48 days | 49-55 days | >55 days |
| --- | --- | --- | --- | --- | --- | --- | --- | --- | --- |
| **LFIA IgG** | 14/34<br>(41%) | 14/31<br>(45%) | 14/18<br>(78%) | 15/19<br>(79%) | 14/18<br>(78%) | 16/25<br>(64%) | 29/38<br>(76%) | 18/32<br>(56%) | 5/7<br>(71%) |
| **LFIA IgM** | 18/34<br>(53%) | 23/31<br>(74%) | 14/18<br>(78%) | 14/19<br>(74%) | 11/18<br>(61%) | 8/25<br>(32%) | 15/38<br>(39%) | 8/32<br>(13%) | 2/7<br>(29%) |
| **LFIA IgG/IgM** | 19/34<br>(56%) | 24/31<br>(77%) | 16/18<br>(89%) | 16/19<br>(84%) | 15/18<br>(83%) | 18/25<br>(72%) | 31/38<br>(82%) | 19/32<br>(59%) | 6/7<br>(86%) |
| **EI ELISA IgG** | 11/23<br>(48%) | 7/19<br>(37%) | 6/6<br>(100%) | 9/10<br>(90%) | 16/17<br>(94%) | 23/26<br>(88%) | 35/39<br>(90%) | 32/38<br>(84%) | 39/40<br>(98%) |
| **EI ELISA IgA** | 11/23<br>(48%) | 12/19<br>(63%) | 6/8<br>(75%) | 9/10<br>(90%) | 14/15<br>(93%) | 21/26<br>(81%) | 32/38<br>(84%) | 29/38<br>(76%) | 13/16<br>(81%) |
| **EDI ELISA IgG** | 24/35<br>(69%) | 24/32<br>(75%) | 20/20<br>(100%) | 28/28<br>(100%) | 27/30<br>(90%) | 28/35<br>(80%) | 36/47<br>(77%) | 26/37<br>(70%) | 14/24<br>(58%) |
| **EDI ELISA IgM** | 21/35<br>(60%) | 17/32<br>(53%) | 19/20<br>(95%) | 11/26<br>(42%) | 13/30<br>(43%) | 9/35<br>(31%) | 15/47<br>(32%) | 6/37<br>(16%) | 3/24<br>(13%) |
| **ECLIA Ig** | 21/34<br>(62%) | 18/26<br>(69%) | 19/19<br>(100%) | 20/21<br>(95%) | 28/29<br>(97%) | 32/34<br>(94%) | 44/45<br>(98%) | 37/39<br>(95%) | 49/53<br>(92%) |
| **CMIA IgG** | 19/31<br>(61%) | 17/28<br>(61%) | 18/18<br>(100%) | 21/22<br>(95%) | 27/28<br>(96%) | 31/32<br>(97%) | 40/43<br>(93%) | 33/36<br>(92%) | 12/13<br>(92%) |
| **LIA IgG** | 17/32<br>(53%) | 14/28<br>(50%) | 16/18<br>(89%) | 18/21<br>(86%) | 26/28<br>(93%) | 27/32<br>(84%) | 42/44<br>(95%) | 32/38<br>(84%) | 24/26<br>(92%) |

**Figure 1.**
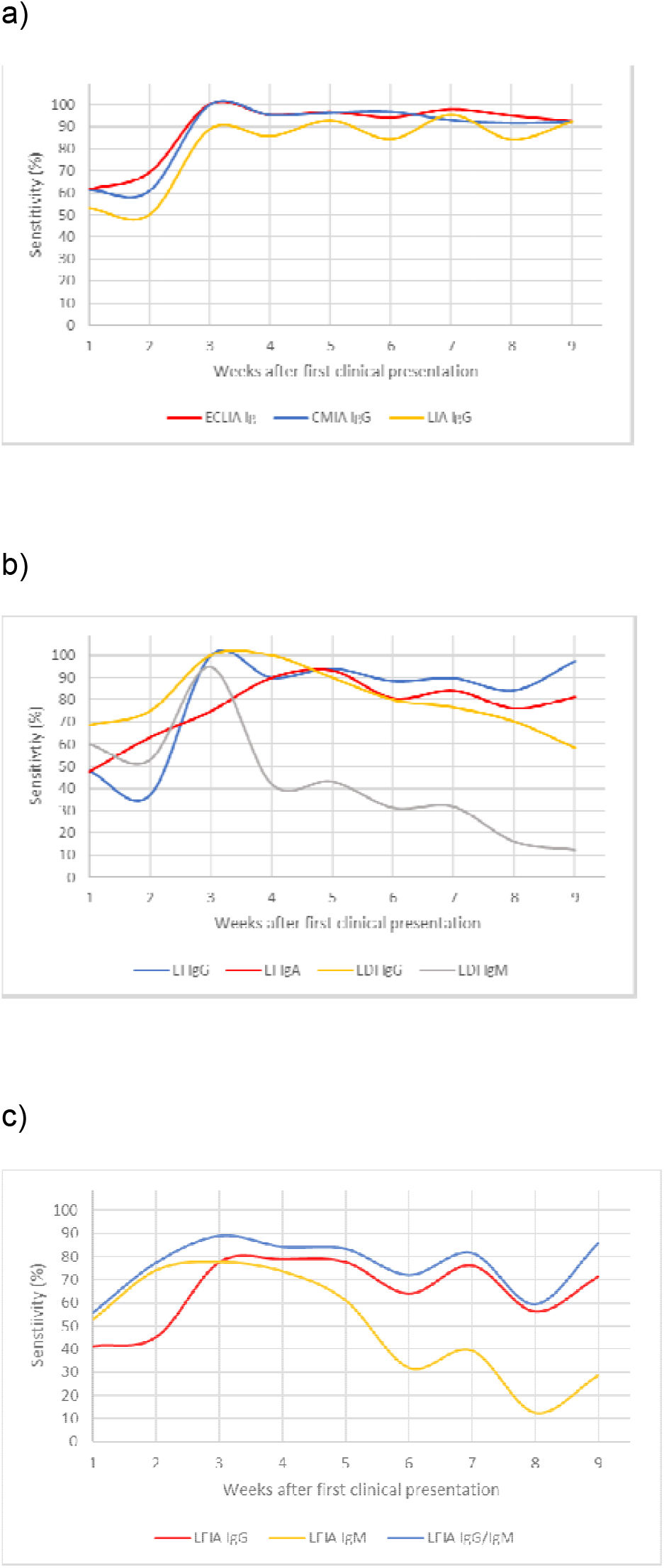
Positivity rates of 10 tests in 8 assays over a period of 9 weeks. a) Chemiluminescence assays, b) ELISAs, and c) LFIA.

### Predictive values of single tests

Predictive values were calculated as a function of pretest probabilities encountered in the clinical routine (i.e., 1% to 40%) to determine the effect of the different diagnostic characteristics at different time points of blood sample collection on the operational test characteristics ^10^. Table 3 illustrates the positive predictive values (PPVs) at different time points, whereas negative predictive values (NPVs) are presented in table 4.

**Table 3.** Heat maps of positive predictive values (PPVs) of different tests used to predict COVID-19 stratified according to the pretest probabilities and sampling time after the first clinical presentation. The predictive values were colored as follows: 99 and 100% in dark blue, 95% to 98% lighter blue, 90% to 94% white, 80% to 89% light yellow, 70% to 79% medium yellow, 60% to 69% dark yellow, 50% to 59% light red, 30% to 49% middle red, and less than 30% dark red.

**CMIA**
| Pretest probability (%) | 0-6 days | 7-13 days | 14-20 days | 21-27 days | 28-34 days | 35-41 days | 42-48 days | 49-55 days | >55 days |
| --- | --- | --- | --- | --- | --- | --- | --- | --- | --- |
| 1 | 55 | 55 | 67 | 66 | 66 | 66 | 65 | 65 | 65 |
| 5 | 87 | 87 | 91 | 91 | 91 | 91 | 91 | 91 | 91 |
| 10 | 93 | 93 | 96 | 96 | 96 | 96 | 95 | 95 | 95 |
| 15 | 96 | 96 | 97 | 97 | 97 | 97 | 97 | 97 | 97 |
| 20 | 97 | 97 | 98 | 98 | 98 | 98 | 98 | 98 | 98 |
| 30 | 98 | 98 | 99 | 99 | 99 | 99 | 99 | 99 | 99 |
| 40 | 99 | 99 | 99 | 99 | 99 | 99 | 99 | 99 | 99 |

**ECLIA**
| Pretest probability (%) | 0-6 days | 7-13 days | 14-20 days | 21-27 days | 28-34 days | 35-41 days | 42-48 days | 49-55 days | >55 days |
| --- | --- | --- | --- | --- | --- | --- | --- | --- | --- |
| 1 | 86 | 87 | 91 | 91 | 91 | 90 | 91 | 91 | 90 |
| 5 | 97 | 97 | 98 | 98 | 98 | 98 | 98 | 98 | 98 |
| 10 | 99 | 99 | 99 | 99 | 99 | 99 | 99 | 99 | 99 |
| 15 | 99 | 99 | 99 | 99 | 99 | 99 | 99 | 99 | 99 |
| 20 | 99 | 99 | 100 | 100 | 100 | 100 | 100 | 100 | 100 |
| 30 | 100 | 100 | 100 | 100 | 100 | 100 | 100 | 100 | 100 |
| 40 | 100 | 100 | 100 | 100 | 100 | 100 | 100 | 100 | 100 |

**LIA**
| Pretest probability (%) | 0-6 days | 7-13 days | 14-20 days | 21-27 days | 28-34 days | 35-41 days | 42-48 days | 49-55 days | >55 days |
| --- | --- | --- | --- | --- | --- | --- | --- | --- | --- |
| 1 | 64 | 63 | 75 | 74 | 76 | 74 | 76 | 74 | 76 |
| 5 | 90 | 90 | 94 | 94 | 94 | 94 | 94 | 94 | 94 |
| 10 | 95 | 95 | 97 | 97 | 97 | 97 | 97 | 97 | 97 |
| 15 | 97 | 97 | 98 | 98 | 98 | 98 | 98 | 98 | 98 |
| 20 | 98 | 98 | 99 | 99 | 99 | 99 | 99 | 99 | 99 |
| 30 | 99 | 99 | 99 | 99 | 99 | 99 | 99 | 99 | 99 |
| 40 | 99 | 99 | 99 | 99 | 100 | 99 | 100 | 99 | 100 |

EI IgG ELISA
| Pretest probability (%) | 0-6 days | 7-13 days | 14-20 days | 21-27 days | 28-34 days | 35-41 days | 42-48 days | 49-55 days | >55 days |
| --- | --- | --- | --- | --- | --- | --- | --- | --- | --- |
| 1 | 38 | 32 | 56 | 53 | 54 | 53 | 53 | 52 | 55 |
| 5 | 76 | 71 | 87 | 86 | 86 | 85 | 86 | 85 | 87 |
| 10 | 87 | 84 | 93 | 93 | 93 | 92 | 93 | 92 | 93 |
| 15 | 91 | 89 | 96 | 95 | 95 | 95 | 95 | 95 | 96 |
| 20 | 94 | 92 | 97 | 97 | 97 | 97 | 97 | 96 | 97 |
| 30 | 96 | 95 | 98 | 98 | 98 | 98 | 98 | 98 | 98 |
| 40 | 98 | 97 | 99 | 99 | 99 | 99 | 99 | 99 | 99 |

EI IgA ELISA
| Pretest probability (%) | 0-6 days | 7-13 days | 14-20 days | 21-27 days | 28-34 days | 35-41 days | 42-48 days | 49-55 days | >55 days |
| --- | --- | --- | --- | --- | --- | --- | --- | --- | --- |
| 1 | 38 | 44 | 49 | 53 | 54 | 51 | 52 | 49 | 51 |
| 5 | 76 | 81 | 83 | 86 | 86 | 84 | 85 | 83 | 84 |
| 10 | 87 | 90 | 91 | 93 | 93 | 92 | 92 | 91 | 92 |
| 15 | 91 | 93 | 94 | 95 | 95 | 95 | 95 | 94 | 95 |
| 20 | 94 | 95 | 96 | 97 | 97 | 96 | 96 | 96 | 96 |
| 30 | 96 | 97 | 98 | 98 | 98 | 98 | 98 | 98 | 98 |
| 40 | 98 | 98 | 98 | 99 | 99 | 99 | 99 | 98 | 99 |

EDI IgG ELISA
| Pretest probability (%) | 0-6 days | 7-13 days | 14-20 days | 21-27 days | 28-34 days | 35-41 days | 42-48 days | 49-55 days | >55 days |
| --- | --- | --- | --- | --- | --- | --- | --- | --- | --- |
| 1 | 14 | 15 | 19 | 19 | 18 | 16 | 16 | 14 | 12 |
| 5 | 46 | 48 | 56 | 56 | 53 | 50 | 49 | 47 | 42 |
| 10 | 64 | 66 | 73 | 73 | 70 | 68 | 67 | 65 | 61 |
| 15 | 74 | 76 | 81 | 81 | 79 | 77 | 76 | 75 | 71 |
| 20 | 80 | 82 | 86 | 86 | 84 | 83 | 82 | 81 | 78 |
| 30 | 88 | 88 | 91 | 91 | 90 | 89 | 89 | 88 | 86 |
| 40 | 92 | 92 | 94 | 94 | 93 | 93 | 92 | 92 | 90 |

EDI IgM ELISA
| Pretest probability (%) | 0-6 days | 7-13 days | 14-20 days | 21-27 days | 28-34 days | 35-41 days | 42-48 days | 49-55 days | >55 days |
| --- | --- | --- | --- | --- | --- | --- | --- | --- | --- |
| 1 | 16 | 14 | 23 | 11 | 12 | 9 | 9 | 5 | 4 |
| 5 | 49 | 46 | 60 | 40 | 41 | 33 | 34 | 21 | 17 |
| 10 | 67 | 64 | 76 | 59 | 59 | 51 | 52 | 35 | 30 |
| 15 | 76 | 74 | 84 | 69 | 70 | 63 | 63 | 46 | 40 |
| 20 | 82 | 80 | 88 | 76 | 77 | 70 | 71 | 55 | 49 |
| 30 | 89 | 87 | 93 | 85 | 85 | 80 | 81 | 68 | 62 |
| 40 | 92 | 91 | 95 | 90 | 90 | 86 | 87 | 77 | 72 |

LFIA IgG
| Pretest probability (%) | 0-6 days | 7-13 days | 14-20 days | 21-27 days | 28-34 days | 35-41 days | 42-48 days | 49-55 days | >55 days |
| --- | --- | --- | --- | --- | --- | --- | --- | --- | --- |
| 1 | 45 | 48 | 61 | 61 | 61 | 56 | 61 | 53 | 59 |
| 5 | 81 | 83 | 89 | 89 | 89 | 87 | 89 | 86 | 88 |
| 10 | 90 | 91 | 95 | 95 | 95 | 93 | 94 | 93 | 94 |
| 15 | 94 | 94 | 96 | 97 | 96 | 96 | 96 | 95 | 96 |
| 20 | 95 | 96 | 97 | 98 | 97 | 97 | 97 | 97 | 97 |
| 30 | 97 | 97 | 99 | 99 | 99 | 98 | 98 | 98 | 98 |
| 40 | 98 | 98 | 99 | 99 | 99 | 99 | 99 | 99 | 99 |

LFIA IgM
| Pretest probability (%) | 0-6 days | 7-13 days | 14-20 days | 21-27 days | 28-34 days | 35-41 days | 42-48 days | 49-55 days | >55 days |
| --- | --- | --- | --- | --- | --- | --- | --- | --- | --- |
| 1 | 11 | 15 | 15 | 14 | 12 | 7 | 8 | 3 | 6 |
| 5 | 39 | 47 | 48 | 47 | 42 | 28 | 32 | 13 | 25 |
| 10 | 57 | 65 | 66 | 65 | 61 | 45 | 50 | 24 | 42 |
| 15 | 68 | 75 | 76 | 75 | 71 | 56 | 61 | 33 | 53 |
| 20 | 75 | 81 | 82 | 81 | 78 | 65 | 69 | 42 | 62 |
| 30 | 84 | 88 | 88 | 88 | 86 | 76 | 79 | 55 | 74 |
| 40 | 89 | 92 | 92 | 92 | 90 | 83 | 86 | 65 | 81 |

LFIA Ig
| Pretest probability (%) | 0-6 days | 7-13 days | 14-20 days | 21-27 days | 28-34 days | 35-41 days | 42-48 days | 49-55 days | >55 days |
| --- | --- | --- | --- | --- | --- | --- | --- | --- | --- |
| 1 | 11 | 14 | 16 | 15 | 15 | 13 | 15 | 11 | 15 |
| 5 | 38 | 46 | 49 | 48 | 48 | 44 | 47 | 39 | 48 |
| 10 | 56 | 64 | 67 | 66 | 66 | 63 | 65 | 58 | 66 |
| 15 | 67 | 74 | 77 | 76 | 75 | 73 | 75 | 69 | 76 |
| 20 | 74 | 80 | 82 | 81 | 81 | 79 | 81 | 76 | 82 |
| 30 | 83 | 87 | 89 | 88 | 88 | 87 | 88 | 84 | 88 |
| 40 | 89 | 91 | 93 | 92 | 92 | 91 | 92 | 89 | 92 |

**Table 4.** Heat maps of negative predictive values (NPVs) of different tests used to predict COVID-19 stratified according to the pretest probabilities and sampling time after the first clinical presentation. The predictive values were colored as follows: 99 and 100% in dark blue, 95% to 98% lighter blue, 90% to 94% white, 80% to 89% light yellow, 70% to 79% medium yellow, 60% to 69% dark yellow.

CMIA
| Pretest probability (%) | 0-6 days | 7-13 days | 14-20 days | 21-27 days | 28-34 days | 35-41 days | 42-48 days | 49-55 days | >55 days |
| --- | --- | --- | --- | --- | --- | --- | --- | --- | --- |
| 1 | 100 | 100 | 100 | 100 | 100 | 100 | 100 | 100 | 100 |
| 5 | 98 | 98 | 100 | 100 | 100 | 100 | 100 | 100 | 100 |
| 10 | 96 | 96 | 100 | 100 | 100 | 100 | 99 | 99 | 99 |
| 15 | 94 | 93 | 100 | 99 | 99 | 99 | 99 | 99 | 99 |
| 20 | 91 | 91 | 100 | 99 | 99 | 99 | 98 | 98 | 98 |
| 30 | 86 | 86 | 100 | 98 | 98 | 99 | 97 | 97 | 97 |
| 40 | 79 | 79 | 100 | 97 | 98 | 98 | 96 | 95 | 95 |

ECLIA
| Pretest probability (%) | 0-6 days | 7-13 days | 14-20 days | 21-27 days | 28-34 days | 35-41 days | 42-48 days | 49-55 days | >55 days |
| --- | --- | --- | --- | --- | --- | --- | --- | --- | --- |
| 1 | 100 | 100 | 100 | 100 | 100 | 100 | 100 | 100 | 100 |
| 5 | 98 | 98 | 100 | 100 | 100 | 100 | 100 | 100 | 100 |
| 10 | 96 | 97 | 100 | 99 | 100 | 99 | 100 | 99 | 99 |
| 15 | 94 | 95 | 100 | 99 | 99 | 99 | 100 | 99 | 99 |
| 20 | 91 | 93 | 100 | 99 | 99 | 99 | 99 | 99 | 98 |
| 30 | 86 | 88 | 100 | 98 | 99 | 98 | 99 | 98 | 97 |
| 40 | 80 | 83 | 100 | 97 | 98 | 96 | 99 | 97 | 95 |

LIA
| Pretest probability (%) | 0-6 days | 7-13 days | 14-20 days | 21-27 days | 28-34 days | 35-41 days | 42-48 days | 49-55 days | >55 days |
| --- | --- | --- | --- | --- | --- | --- | --- | --- | --- |
| 1 | 100 | 99 | 100 | 100 | 100 | 100 | 100 | 100 | 100 |
| 5 | 98 | 97 | 99 | 99 | 100 | 99 | 100 | 99 | 100 |
| 10 | 95 | 95 | 99 | 98 | 99 | 98 | 100 | 98 | 99 |
| 15 | 92 | 92 | 98 | 98 | 99 | 97 | 99 | 97 | 99 |
| 20 | 89 | 89 | 97 | 97 | 98 | 96 | 99 | 96 | 98 |
| 30 | 83 | 82 | 95 | 94 | 97 | 94 | 98 | 94 | 97 |
| 40 | 76 | 75 | 93 | 91 | 95 | 91 | 97 | 90 | 95 |

EI IgG ELISA
| Pretest probability (%) | 0-6 days | 7-13 days | 14-20 days | 21-27 days | 28-34 days | 35-41 days | 42-48 days | 49-55 days | >55 days |
| --- | --- | --- | --- | --- | --- | --- | --- | --- | --- |
| 1 | 99 | 99 | 100 | 100 | 100 | 100 | 100 | 100 | 100 |
| 5 | 97 | 97 | 100 | 99 | 100 | 99 | 99 | 99 | 100 |
| 10 | 94 | 93 | 100 | 99 | 99 | 99 | 99 | 98 | 100 |
| 15 | 92 | 90 | 100 | 98 | 99 | 98 | 98 | 97 | 100 |
| 20 | 88 | 86 | 100 | 98 | 99 | 97 | 97 | 96 | 99 |
| 30 | 82 | 79 | 100 | 96 | 98 | 95 | 96 | 94 | 99 |
| 40 | 74 | 70 | 100 | 94 | 96 | 93 | 94 | 90 | 98 |

EI IgA ELISA
| Pretest probability (%) | 0-6 days | 7-13 days | 14-20 days | 21-27 days | 28-34 days | 35-41 days | 42-48 days | 49-55 days | >55 days |
| --- | --- | --- | --- | --- | --- | --- | --- | --- | --- |
| 1 | 99 | 100 | 100 | 100 | 100 | 100 | 100 | 100 | 100 |
| 5 | 97 | 98 | 99 | 99 | 100 | 99 | 99 | 99 | 99 |
| 10 | 94 | 96 | 97 | 99 | 99 | 98 | 98 | 97 | 98 |
| 15 | 92 | 94 | 96 | 98 | 99 | 97 | 97 | 96 | 97 |
| 20 | 88 | 92 | 94 | 98 | 98 | 95 | 96 | 94 | 95 |
| 30 | 82 | 86 | 90 | 96 | 97 | 92 | 94 | 91 | 93 |
| 40 | 74 | 80 | 86 | 94 | 96 | 89 | 90 | 86 | 89 |

EDI IgG ELISA
| Pretest probability (%) | 0-6 days | 7-13 days | 14-20 days | 21-27 days | 28-34 days | 35-41 days | 42-48 days | 49-55 days | >55 days |
| --- | --- | --- | --- | --- | --- | --- | --- | --- | --- |
| 1 | 100 | 100 | 100 | 100 | 100 | 100 | 100 | 100 | 100 |
| 5 | 98 | 99 | 100 | 100 | 99 | 99 | 99 | 98 | 98 |
| 10 | 96 | 97 | 100 | 100 | 99 | 98 | 97 | 97 | 95 |
| 15 | 95 | 96 | 100 | 100 | 98 | 96 | 96 | 95 | 93 |
| 20 | 92 | 94 | 100 | 100 | 97 | 95 | 94 | 93 | 90 |
| 30 | 88 | 90 | 100 | 100 | 96 | 92 | 91 | 88 | 84 |
| 40 | 82 | 85 | 100 | 100 | 93 | 88 | 86 | 83 | 78 |

EDI IgM ELISA
| Pretest probability (%) | 0-6 days | 7-13 days | 14-20 days | 21-27 days | 28-34 days | 35-41 days | 42-48 days | 49-55 days | >55 days |
| --- | --- | --- | --- | --- | --- | --- | --- | --- | --- |
| 1 | 100 | 100 | 100 | 99 | 99 | 99 | 99 | 99 | 99 |
| 5 | 98 | 98 | 100 | 97 | 97 | 96 | 96 | 96 | 95 |
| 10 | 96 | 95 | 99 | 94 | 94 | 93 | 93 | 91 | 91 |
| 15 | 93 | 92 | 99 | 90 | 91 | 89 | 89 | 87 | 86 |
| 20 | 91 | 89 | 99 | 87 | 87 | 85 | 85 | 82 | 82 |
| 30 | 85 | 83 | 98 | 80 | 80 | 77 | 77 | 73 | 72 |
| 40 | 78 | 76 | 97 | 72 | 72 | 68 | 68 | 63 | 62 |

LFIA IgG
| Pretest probability (%) | 0-6 days | 7-13 days | 14-20 days | 21-27 days | 28-34 days | 35-41 days | 42-48 days | 49-55 days | >55 days |
| --- | --- | --- | --- | --- | --- | --- | --- | --- | --- |
| 1 | 99 | 99 | 100 | 100 | 100 | 100 | 100 | 100 | 100 |
| 5 | 97 | 97 | 99 | 99 | 99 | 98 | 99 | 98 | 99 |
| 10 | 94 | 94 | 98 | 98 | 98 | 96 | 97 | 95 | 97 |
| 15 | 91 | 91 | 96 | 96 | 96 | 94 | 96 | 93 | 95 |
| 20 | 87 | 88 | 95 | 95 | 95 | 92 | 94 | 90 | 93 |
| 30 | 80 | 81 | 91 | 92 | 91 | 87 | 91 | 84 | 89 |
| 40 | 72 | 73 | 87 | 88 | 87 | 81 | 86 | 77 | 84 |

LFIA IgM
| Pretest probability (%) | 0-6 days | 7-13 days | 14-20 days | 21-27 days | 28-34 days | 35-41 days | 42-48 days | 49-55 days | >55 days |
| --- | --- | --- | --- | --- | --- | --- | --- | --- | --- |
| 1 | 100 | 100 | 100 | 100 | 100 | 99 | 99 | 99 | 99 |
| 5 | 97 | 99 | 99 | 99 | 98 | 96 | 97 | 95 | 96 |
| 10 | 95 | 97 | 97 | 97 | 96 | 93 | 93 | 91 | 92 |
| 15 | 92 | 95 | 96 | 95 | 93 | 89 | 90 | 86 | 88 |
| 20 | 89 | 94 | 95 | 94 | 91 | 85 | 86 | 81 | 84 |
| 30 | 83 | 90 | 91 | 89 | 85 | 77 | 79 | 72 | 76 |
| 40 | 75 | 85 | 87 | 85 | 79 | 68 | 70 | 62 | 67 |

LFIA Ig
| Pretest probability (%) | 0-6 days | 7-13 days | 14-20 days | 21-27 days | 28-34 days | 35-41 days | 42-48 days | 49-55 days | >55 days |
| --- | --- | --- | --- | --- | --- | --- | --- | --- | --- |
| 1 | 100 | 100 | 100 | 100 | 100 | 100 | 100 | 100 | 100 |
| 5 | 98 | 99 | 99 | 99 | 99 | 98 | 99 | 98 | 99 |
| 10 | 95 | 97 | 99 | 98 | 98 | 97 | 98 | 95 | 98 |
| 15 | 92 | 96 | 98 | 97 | 97 | 95 | 97 | 93 | 97 |
| 20 | 90 | 94 | 97 | 96 | 96 | 93 | 95 | 90 | 96 |
| 30 | 83 | 91 | 95 | 93 | 93 | 89 | 92 | 85 | 94 |
| 40 | 76 | 86 | 93 | 90 | 90 | 84 | 89 | 78 | 91 |

The chemiluminescence assays displayed good predictive characteristics in excluding COVID-19 with a negative test result if the sample was collected at week 3 or thereafter. In situations with a moderate to high pretest probability, none of the chemiluminescence assays would be capable of excluding a COVID-19 infection during the first two weeks of a suspected infection. A positive test result obtained using CMIA and LIA for asymptomatic patients with a very low and low pretest probability had a PPV of less than 95%, regardless of the time point at which the blood sample was collected. Positive results obtained using the ECLIA at various sampling time points from patients with a low pretest probability of at least 5% had a PPV ≥95% for the prediction of COVID-19.

Regarding the ELISAs, the operational test characteristics of the assay from one manufacturer are better than the assay from another manufacturer. The EDI IgG and IgM ELISAs, however, generally showed low PPVs during the first two weeks and thereafter in low pretest probability settings, independent of the blood sampling time point. The ability of the EI ELISAs to exclude COVID-19 during the first two weeks with a NPV of 95% or greater is only possible only in patients with very low and low pretest probabilities. Thereafter, the exclusion of COVID-19 with a negative test result was able to be relatively safely conducted in samples from patients with higher pretest probabilities. The EDI IgG ELISA excluded COVID-19 with NPVs ≥95% for patients with very low, low and moderate pretest probabilities, whereas the IgM assay cannot be reliably used in this context. Regarding a positive diagnosis of COVID-19, neither EDI ELISAs appeared to add valuable information in any situation when used as single tests.

The LFIA IgG measurement appeared to possess the potential for diagnosing COVID-19 in patients with moderate and higher pretest probabilities, but not in patients with very low and low pretest probabilities, when blood samples were collected at least 3 weeks after symptom onset. Positive LFIA IgM results did not display a useful PPV in predicting COVID-19. The combined judgment of both IgG and IgM LFIA tests was not useful in reliably predicting COVID-19. The combined LFIA test possessed an NPV of ≥95% in excluding COVID-19 in patients with very low and low pretest probabilities (i.e., symptomatic and asymptomatic patients), regardless of the time of sampling. Thereafter, the NPV also reached values of ≥95% for patients with moderate pretest probabilities. Together, the LFIA appeared to exclude COVID-19 in patients with low pretest probabilities throughout the disease course with negative combined IgG/IgM results. Furthermore, LFIA IgG tests also excluded COVID-19 in patients with a moderate pretest probability at week 3 after the first clinical presentation and thereafter.

### Orthogonal testing algorithms

In addition to the IgM ELISA, all investigated tests provided relatively high NPVs in excluding COVID-19 in patients with a negative test results and low to moderate pretest probabilities over the whole 9 weeks. With the exception of the ECLIA, the PPVs were less than 95% for individuals with low pretest probabilities over the whole 9 weeks. Chemiluminescence tests for patients with a moderate pretest probability provided PPVs ≥95%, whereas the PPVs for ELISAs and LFIA were lower. We therefore determined the PPVs using an orthogonal testing approach, where positive results were confirmed with another independent test. The combination of two of the lowest performing tests for positive results in an orthogonal testing algorithm, i.e., the LFIA IgG/IgM test followed by the EDI IgG ELISA test, for patients with a moderate pretest probability consistently provided PPVs of ≥95%, as shown in table 5. A combination of two chemiluminescence tests (i.e., CMIA followed by LIA) provided PPVs of 100% for patients with all pretest probabilities (table 5). Other test combinations (i.e., ECLIA/EDI IgG, ECLIA/LIA, ECLIA/CMIA, ECLIA/EI IgG, CMIA/EDI IgG, CMIA/EI IgG, LIA/EDI IgG, and LIA/EI IgG) provided comparable PPVs (≥99%) to the CMIA/LIA combination throughout the whole 9 week period for patients with all pretest probabilities (data not shown). NPVs of a CMIA/LIA combination were lower than the CMIA or the LIA alone, as shown in table 6. Other combinations of negative test results exhibited similar performance and are not shown.

**Table 5.**
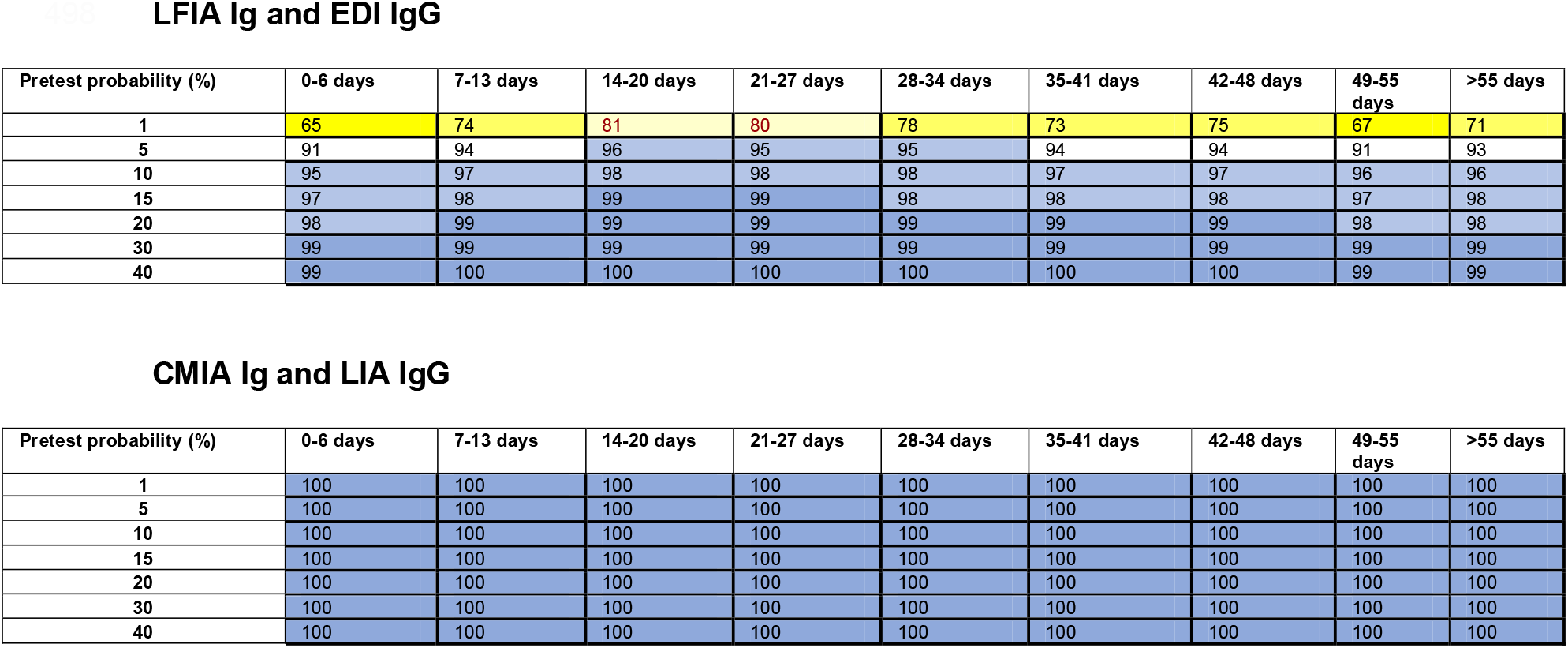
PPVs obtained for the combinations of two assays performed according to the orthogonal testing algorithm recommended by the FDA and CDC. The predictive values were colored as follows: 99 and 100% in dark blue, 95% to 98% lighter blue, 90% to 94% white, 80% to 89% light yellow, 70% to 79% medium yellow, 60% to 69% dark yellow.

**Table 6.** NPVs obtained for the combination of the CMIA and LIA for samples with an initial negative test result using an orthogonal test algorithm. NPVs of the test combination are lower than either single test. The predictive values were colored as follows: 90% to 94% white, 80% to 89% light yellow, 70% to 79% medium yellow, 60% to 69% dark yellow, 50% to 59% light red, 30% to 49% middle red, and less than 30% dark red.

CMIA Ig and LIA IgG
| Pretest probability (%) | 0-6 days | 7-13 days | 14-20 days | 21-27 days | 28-34 days | 35-41 days | 42-48 days | 49-55 days | >55 days |
| --- | --- | --- | --- | --- | --- | --- | --- | --- | --- |
| 1 | 63 | 62 | 82 | 78 | 88 | 77 | 92 | 77 | 87 |
| 5 | 25 | 24 | 46 | 41 | 58 | 39 | 69 | 40 | 57 |
| 10 | 14 | 13 | 29 | 25 | 40 | 23 | 52 | 24 | 39 |
| 15 | 9 | 9 | 20 | 17 | 29 | 16 | 40 | 16 | 28 |
| 20 | 7 | 6 | 15 | 13 | 23 | 12 | 32 | 12 | 22 |
| 30 | 4 | 4 | 10 | 8 | 15 | 7 | 26 | 7 | 14 |
| 40 | 3 | 2 | 6 | 5 | 10 | 5 | 15 | 5 | 10 |

## Discussion

A recent Cochrane review stated that data available for diagnostic characteristics of SARS-CoV-2 antibody assays for more than 35 days after symptom onset are insufficient ^3^. The present study provides data for a variety of tests beyond 9 weeks (63 days). We confirm that the investigated tests have insufficient test characteristics, particularly for positive results, two weeks after the first clinical presentation, with one exception. The combination of tests in an orthogonal testing algorithm, even tests with suboptimal test characteristics, for positive results provided PPVs ≥95% for individuals with a moderate and high pretest probabilities early in the course of the infection. Furthermore, positivity rates appeared to be stable for chemiluminescence assays testing for total Ig and IgG, whereas the positivity rates of IgM tests decreased over time.

Fenwick and colleagues and others observed sensitivities than the present study in the ECLIA (95.6%), LIA (88.9%), EI IgG (88.9%), and EDI IgG (76.7%), which corroborates our findings derived in smaller cohorts stratified according to time after first clinical presentation of COVID-19 ^15-18^. The sensitivity of the CMIA described by Tang et al. (93.8%) is comparable to the sensitivity values observed in our study ^18^. The sensitivity of the LFIA was also comparable to the values reported for other LFIA test formats (75% in the second week and 90% thereafter) ^19^. Some test formats analyzed in our study had stable sensitivities over a duration of 9 weeks after the first clinical presentation. Other test formats, particularly tests measuring IgM and IgA, as well as an ELISA measuring IgG directed against the nucleocapsid antigen, showed decreasing positivity rates over time. We consider the use of test formats with decreasing sensitivities over time problematic, as long as they do not employ modified cut-off values, which should be further investigated in larger cohorts.

In clinical diagnostics, the operational characteristics rather than the diagnostic characteristics are of interest. In our opinion, the use of heat maps to illustrate diagnostic strengths and weaknesses in certain situations according to the time since the first clinical presentation and pretest probability is useful, particularly when only one test is employed. To the best of our knowledge, heat maps have not yet been introduced as a tool to interpret serological assays of infectious diseases ^4^. Unfortunately, at the moment, there is no clinical score currently unavailable to assess pretest probability, which relies on a history of clinical symptoms alone and could also be utilized retrospectively (e.g., because no laboratory results are available at the time when a patient was symptomatic) ^20^. However, other rough estimates also allow for assessment of pretest probability and clinical use of the heatmaps according to the individual situation of a patient: symptomatic patients have a pretest probability of 10% and higher, asymptomatic patients usually have a pretest probability of less than 10%, close contacts of patients with confirmed COVID-19 cases have a pretest probability of 15-30% ^10^. Seroprevalence data of a region can also help to assess the pretest probability even in the absence of clinical symptoms.

Positive predictive values differed among the different test formats. Based on the results of our study, chemiluminescence formats have somewhat better operative characteristics than at least some of the investigated ELISA and LFIA test formats. Furthermore, the SARS-CoV-2 IgM test should be used with caution to assess an immune response to COVID-19 that has already been apparent for longer than 21 days. This result is consistent with the findings of Costa et al, who, because of the simultaneous occurrence with IgG, judged IgM to be of no value in diagnosing acute or subacute SARS-CoV-2 infections ^21^. Our study clearly illustrates that an orthogonal testing approach adds valuable diagnostic information. The combination of two of the lowest performing assays according to the approach recommended by FDA and CDC for moderate pretest probabilities provided PPVs ≥95% throughout the 9 weeks after the first clinical presentation. These characteristics further improved and were extended to low prevalence settings when the chemiluminescence and EI IgG assays were combined. Our study confirms that an orthogonal testing approach for initially positive samples adds diagnostic value.

After two weeks, negative predictive values of tests investigating IgG or total Ig were ≥ 95% in patients with very low, low and moderate probabilities. Chemiluminescence assays displayed a somewhat higher predictivity than ELISAs and LFIA. Importantly, the LFIA reading for both IgM and IgG bands showed greater predictive value than reading either of the two bands alone. However, in contrast to the practice of confirming positive test results with an alternative test, the application of an orthogonal testing approach is not useful in a sample with an initially negative test result. NPVs were worse than the initial index test.

Our study has several strengths and limitations. One strength is that we were able to study positivity rates over 9 weeks. Another strength is that we studied 8 different assays employing the commonly used measurement principles, i.e., chemiluminescence, ELISA, and lateral flow immunoassay. A limitation represents is that the day of onset of clinical symptoms was not available. Instead, we recorded the first date of clinical presentation for COVID-19. In Liechtenstein and Switzerland, all symptomatic patients are tested for COVID-19 using PCR. Due to the high accessibility of testing in COVID-19 testing centers, patients presented to these centers very rapidly after the onset of symptoms. Additionally, our data, particularly the results obtained within the first 35 days, the findings described elsewhere using the onset of symptoms as the reference time point ^3,18,22^. Some studies have also presented data using PCR testing as a reference time point ^18,22^. We therefore propose that our approach of using the first clinical presentation for COVID-19 testing in the investigated setting is a good proxy for the onset of symptoms. When analyzing the different time strata, the sample size of the included samples was not large, and not all sera were tested with all tests, which might result in relatively imprecise point estimates of sensitivity. The sample sizes are comparable to the values reported in other studies, and the sensitivities identified in our setting are comparable to the values reported in larger other studies, suggesting that relatively small sample sizes do not invalidate our findings ^3,15,17,18,21,22^. Finally, even if we did not have a detailed clinical description of the patients, we know, that the samples of our patients originated from outpatient settings. Thus it can be assumed, that included confirmed symptomatic patients with mild to moderate COVID-19 severity. Our findings might not apply to individuals with severe cases and an asymptomatic disease course ^15,23^, which requires confirmation in further investigations. Our findings, however, provide valuable insights into interpreting past symptoms in patients who were not assessed using RT-PCR testing or who had a negative RT-PCR test, despite the presence of suggestive symptoms, at least within two months of clinical presentation.

In conclusion, we identified sensitivities for 8 different SARS-CoV-2 antibody tests in symptomatic patients over a period of 63 days. The investigated chemiluminescence assays did not exhibit a decrease in positivity rates over the investigated period. This finding was not observed for other test formats, e.g., IgM tested using ELISA or LFIA. Not all commercially available and CE-marked assays exhibit satisfactory predictive values for diagnosing or excluding COVID-19. In particular, predictive values were lower during the first two weeks after first clinical presentation. An orthogonal test algorithm confirming positive, but not negative, results from a single test with an independent assay provided satisfactory positive predictive values, even for tests with less accurate performance. Orthogonal testing approaches for positive results should therefore be reinforced.

## Data Availability

The data that support the findings of this study are available from the corresponding author, [LR], upon reasonable request.

## Acknowledgments

The assistance of Toni Schönenberger and Walter Frehner in identifying samples is acknowledged.

## Author credit statement

**Martin Risch:** Conceptualization, Methodology, Formal analysis, and Writing - Original Draft; **Myriam Weber:** Conceptualization, Methodology, and Formal analysis; **Sarah Thiel:** Data curation, Validation, and Resources; **Kirsten Grossmann:** Resources; **Kirsten Nadja Wohlwend**: Investigation, Validation, and Resources; **Thomas Lung:** Validation and Resources; **Dorothea Hillmann:** Investigation, Validation, Project administration, and Resources; **Michael Ritzler:** Resources; **Francesca Ferrara:** Resources; **Susanna Bigler:** Validation and Resources; **Konrad Egli:** Validation and Resources; **Thomas Bodmer:** Validation and Resources; **Mauro Imperiali:** Validation and Resources; **Yacir Salimi:** Validation and Resources; **Felix Fleisch:** Resources; **Alexia Cusini:** Resources; **Harald Renz:** Conceptualization; **Philipe Kohler:** Validation and Writing - Original Draft; **Pietro Vernazza:** Conceptualization and Writing - Original Draft; **Christian Kahlert:** Conceptualization, Methodology Supervision, Resources, and Writing - Original Draft; **Matthias Paprotny:** Supervision and Resources; **Lorenz Risch:** Conceptualization, Methodology, Funding acquisition, Supervision, Resources, and Writing - Original Draft; **All authors:** Writing - Reviewing and Editing

